# Low Frequency Oscillations in the Medial Orbitofrontal Cortex Mediate Widespread Hyperalgesia Across Pain Conditions

**DOI:** 10.1101/2025.06.15.25329637

**Authors:** Hyung G. Park, George Kenefati, Mika M. Rockholt, Xiaomeng Ju, Rachel R. Wu, Zhe Sage Chen, Tamas A. Gonda, Jing Wang, Lisa V. Doan

**Author notes:** Correspondence: Hyung G. Park, PhD, Jing Wang, MD, PhD, Lisa V. Doan, MD.

## Abstract

Widespread hyperalgesia, characterized by pain sensitivity beyond the primary pain site, is common across chronic pain conditions, yet it is unknown if there are shared neural mechanisms that can function as a biosignature for this condition. We used high-density electroencephalography (EEG) to examine noxious stimulus-evoked cortical activity at both disease and disease-free sites in patients with chronic low back pain (cLBP) and chronic pancreatitis (CP). Compared with healthy controls and patients with localized pain, cLBP patients with widespread hyperalgesia exhibited altered delta, theta, and alpha oscillations in the bilateral medial orbitofrontal cortex (mOFC). To test if this neural signature for widespread hyperalgesia is shared across pain conditions, we applied this mOFC-derived neural signature to an independent cohort of CP patients. We found that this signature not only successfully distinguished CP patients with widespread hyperalgesia but also predicted their pain response to peripherally-targeted interventions. These findings suggest that altered mOFC oscillatory responses may reflect a shared neural mechanism of widespread hyperalgesia across chronic pain conditions.

## Introduction

Widespread hyperalgesia is characterized by increased sensitivity to peripheral noxious stimuli in a diffuse, non-anatomic distribution and is considered a marker of impaired central pain processing and nociplastic pain^1–4^. Widespread hyperalgesia can be found in many chronic pain syndromes, including chronic low back pain (cLBP), chronic pancreatitis (CP), fibromyalgia, knee osteoarthritis, and rheumatoid arthritis^5–10^. It is associated with high levels of pain severity and poor response to peripherally-directed treatments^11, 12^. Despite its high prevalence, mechanisms of widespread hyperalgesia remain poorly understood, and while the central nervous system is thought to play a key role, it is unclear if unifying brain mechanisms are shared across multiple pain syndromes.

Neuroimaging studies of the brain have shown structural and functional alterations in chronic pain^13^. Key regions involved in the top-down aversive and cognitive processing of pain include the medial orbitofrontal cortex (mOFC)^10^ and prefrontal cortex (PFC)^6, 14^. Recently, intracranial recordings in humans with chronic neuropathic pain showed that spontaneous pain severity was decoded by delta power in the mOFC^14^. Meanwhile, resting state magnetoencephalography recordings in patients with fibromyalgia, a prototypical nociplastic pain condition, also revealed that changes in theta, beta, and gamma power in the mOFC and other prefrontal areas are associated with higher affective pain scores^15^.

Advances in source localization have enabled high density (>64 channels) electroencephalogram (EEG) recordings to provide anatomical information to facilitate functional studies^16–18^. For example, amplitudes of beta and gamma oscillations in prefrontal areas were shown to positively correlate with ongoing pain^19,20^. Abnormal alpha oscillations also play a role in chronic pain^21–23^, as the amplitude and timing of peak alpha frequency have been correlated with pain intensity^24–27^. Resting state EEG and functional MRI (fMRI) recordings can provide data regarding chronic alterations in brain circuits in the absence of sensory inputs^28^. Meanwhile, combining highly temporally specific EEG recordings with quantitative sensory testing (QST) that introduces noxious stimuli can assess how the presence of chronic pain alters nociceptive processing^6, 16, 29^, to provide insights into mechanisms underlying widespread hyperalgesia^30–33^.

In a prior study assessing EEG responses to acute nociceptive stimuli in cLBP participants, we found that altered theta and alpha oscillations in the mOFC and the dorsolateral PFC (dlPFC) were associated with hypersensitivity on QST^6^. A subset of cLBP participants experienced widespread hyperalgesia manifested as hypersensitivity at locations unrelated to the diseased site. However, it is unclear whether stimulus-evoked brain oscillatory activities seen in cLBP patients with widespread hyperalgesia will differ from those seen in patients with localized pain in response to nociceptive input. An even more interesting question is whether specific brain oscillatory findings associated with widespread hyperalgesia in cLBP participants will generalize to widespread hyperalgesia in other chronic pain conditions.

To test whether there are shared brain mechanisms that can function as potential biosignatures for widespread hyperalgesia across different pain conditions, we studied oscillatory activities using high-density (64-ch) EEG recordings during QST at both diseased and disease-free sites in cLBP patients. We focused our inquiry on the mOFC and dlPFC, given their prominent roles in chronic pain phenotypes. We found increased delta and theta oscillations and decreased alpha oscillations in the bilateral mOFC were associated with widespread hyperalgesia in cLBP participants. To test whether these cortical changes reflect a shared mechanism underlying widespread hyperalgesia, we applied this oscillatory brain signature to evoked EEG responses in participants with painful CP. We found that the cLBP-derived EEG signature successfully distinguished widespread hyperalgesia in CP participants. Furthermore, we showed that this biosignature, when evaluated at baseline, successfully predicted pain response in CP patients undergoing endoscopic retrograde pancreatography (ERP) for ductal obstruction, a peripherally-directed treatment that is unlikely to alleviate centrally-mediated pain. These results suggest that disrupted nociceptive response in the mOFC likely constitutes a common mechanism for widespread hyperalgesia that has the potential to predict treatment responses for chronic pain.

## Methods

We analyzed data from 85 pooled sessions of QST and simultaneous high-density EEG recordings, obtained from two prospective cohort studies involving participants with cLBP and CP. Both studies were approved by the NYU Grossman School of Medicine IRB (8/22/2019, #i19-01088; 9/18/2023, #i23-00766) and conducted in accordance with the Declaration of Helsinki. All participants provided written informed consent.

### Study Participants and Eligibility Criteria

Data were collected from two cohorts: 38 cLBP participants (including 4 with long-term longitudinal follow-up; n = 42) vs. 25 pain-free controls, and 18 participants with painful CP. cLBP inclusion criteria included adults aged 18 to 75 years with pain lasting greater than 6 months and baseline average pain intensity > 4 on a 0 to10 point numerical rating scale (NRS), with exclusion criteria including acute lumbosacral radiculopathy with motor or sensory loss, systemic symptoms, history of cognitive impairment or clinical signs of altered mental status, history of schizophrenia, daily benzodiazepine use, and pregnancy.

Inclusion criteria for pain-free healthy controls included age 18 to 75 years and American Society of Anesthesiologists (ASA) physical status 1-3. CP inclusion criteria included adults older than 18 years with Cambridge III-IV or M-ANNHEIM criteria for definitive chronic pancreatitis^34, 35^, pain present ≥ 3 days per week for ≥ 3 months with average pain ≥ 4 on the 0 to 10 point NRS and scheduled for endoscopic therapy for ductal obstruction. Exclusion criteria included recent acute pancreatitis within 2 months of enrollment, technically successful ERP in 3 months prior to enrollment, active illicit drug use (excluding marijuana) or chronic benzodiazepine use, ASA physical status > 3, immune-mediated pancreatitis, pancreatic neoplasms, major neurological disease, and pregnancy. At baseline, pain was assessed in the cLBP cohort with the PROMIS pain intensity 1a questionnaire which assesses average pain in the prior week. In the CP cohort, pain was assessed with the Brief Pain Inventory-short form (BPI) pain severity subscale^36^.

### EEG Recordings

EEG data were recorded using a 64-channel cap (G.Tec or Compumedics Neuroscan) equipped with two integrated bipolar leads for vertical electrooculogram with the ground electrode positioned on the left cheek, sampled at 1000 Hz and filtered between 0.5–100 Hz. Each recording session began with baseline recordings with 5 minutes eyes closed and 5 minutes eyes opened. For each participant, stimulus-evoked EEG was recorded during mechanical pinprick stimuli at four stimulus conditions (see **QST Protocols**). See **Figure 1** for the experiment scheme.

**Figure 1.**
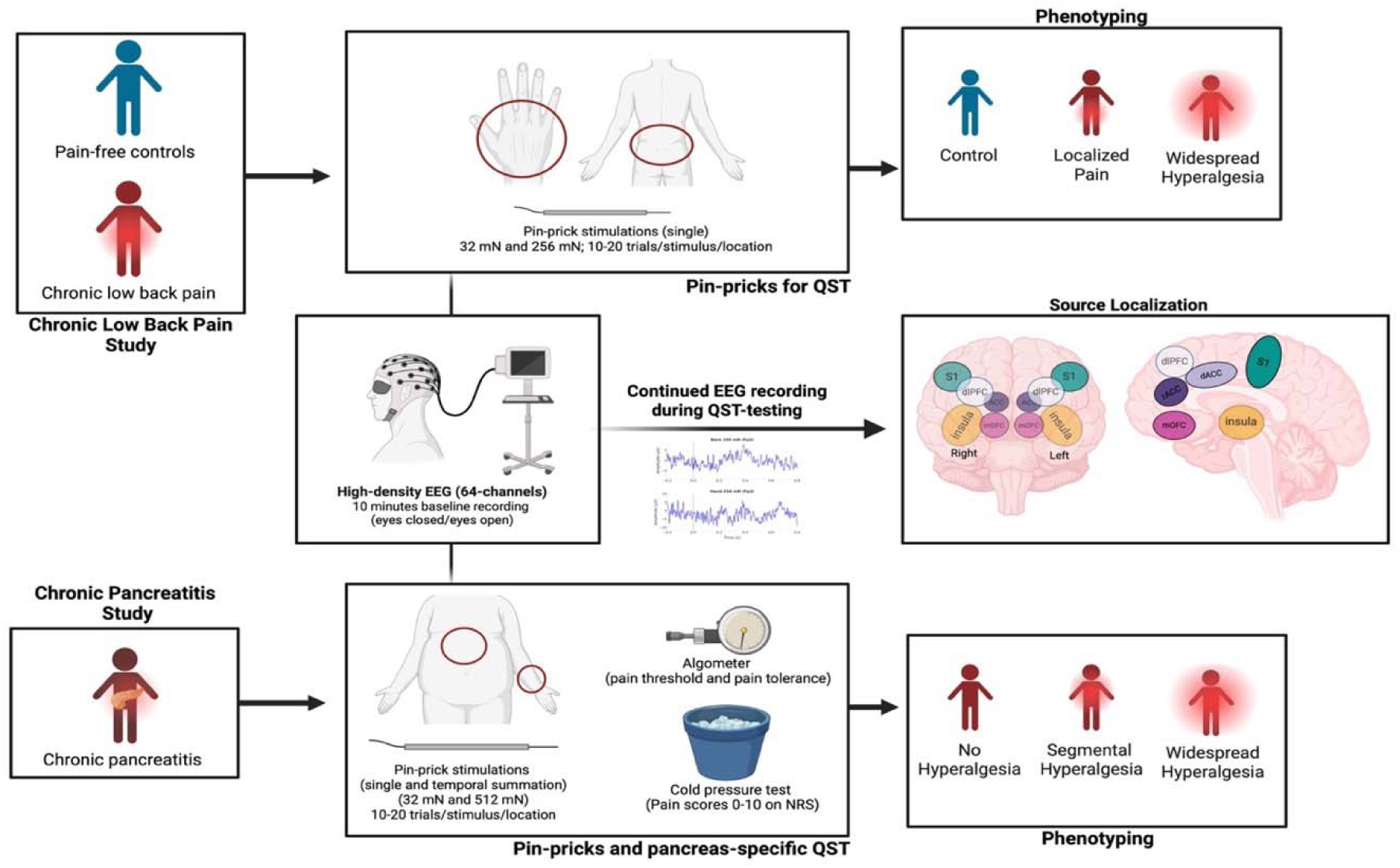
**Schematic overview of the experimental design for concurrent QST and EEG**. **Left panels**: Participants with chronic low back pain (cLBP) and chronic pancreatitis (CP) were enrolled. **Middle panels**: EEG was recorded during pinprick stimulation at the lower back and dorsum of the right hand (for cLBP), and at the upper abdomen and dominant forearm (for CP). **Right panels**: Participants were pain-phenotyped, and EEG source localization was performed, focusing on mOFC and dlPFC as regions of interest (ROIs).

### QST Protocols

In the cLBP cohort (n = 67), participants (with cLBP and pain-free healthy controls) were blindfolded and instructed to remain relaxed but awake during EEG recordings. Mechanical pinprick stimuli (MRC Systems GmbH, Heidelberg, Germany) were applied to the lower back (pain affected site for cLBP participants) and the dorsum of the right hand (site typically unaffected by pain) using forces of 32 mN and 256 mN. Each force was delivered in 10-20 randomized trials per site with an interstimulus interval of approximately 10 seconds, and participants rated each stimulus on the 0 to 10 point NRS scale.

Analogous to the cLBP cohort, the CP cohort (n=18) underwent a stimulus-evoked EEG protocol. Mechanical pinprick stimuli of 32 mN and 512 mN were applied to the upper abdomen (pain affected site in CP) and the dominant forearm (site typically unaffected by pain in CP). Participants rated each stimulus on the NRS scale. CP participants also underwent a standardized pancreatic QST (P-QST) protocol^8, 37^. Briefly, participants received repetitive pinprick stimulation with a 256 mN force to assess temporal summation at both the upper abdomen and the midline volar forearm of the dominant arm. CP participants underwent muscle pressure testing using an algometer (Algometer Type II, SBMEDIC Electronics, Solna, Sweden) to determine pressure pain detection thresholds (PDT) and pain tolerance thresholds (PTT). Pressure was applied to the following dermatomes: C5 (clavicle), T10 (upper epigastric area—pancreatic viscerotome), T10 back (posterior pancreatic viscerotome), L1 (anterior superior iliac crest), and L4 (quadriceps). Lastly, the conditioned pain modulation (CPM) was assessed with a cold pressor test, in which the dominant hand was immersed in ice-chilled water for up to 120 seconds, and PTT at the non-dominant quadriceps muscle, measured before and after the cold pressor test.

### Pain Phenotyping

For the cLBP cohort, a threshold for hyperalgesic response to 32 mN stimulation at the hand (a site typically unaffected by pain) was defined as 2 standard deviations above the mean pain rating in control participants^38^. cLBP participants below this threshold were classified as having localized Pain, whereas those above the threshold were classified as having widespread hyperalgesia. Control participants were classified as no pain.

For the CP participants, pain phenotypes were determined using the published algorithm for the P-QST protocol^8, 37^ defined based on cold pressor test, CPM, PDT, and temporal summation distinguishing the participants into three groups: no hyperalgesia, segmental hyperalgesia, and widespread hyperalgesia.

### Standardized ERP in CP Cohort and Pain Outcome

As part of routine standard of care, CP participants underwent standardized ERP. A technically successful procedure was defined as stent placement across a pancreatic duct stricture and/or removal of obstructing stone or intraductal calcification. BPI was assessed at baseline before ERP and at approximately 3 months after a technically successful procedure.

### EEG Preprocessing

Preprocessing included notch filtering (at 60 Hz with 3 Hz notch width) to remove powerline noise, band-pass filtering, bad channel detection and interpolation using the PyPREP toolbox, and artifact correction via independent component analysis (ICA)^39–41^. Source localization was performed using the MNE-Python toolbox with the minimum norm estimate (MNE) method, employing a boundary-element head model and Desikan-Killiany atlas. Time-frequency representations were computed using the mne.time_frequency.tfr_multitapter function from the MNE-Python toolbox with default parameters^42^. EEG data were epoched from-1 to 1 second relative to stimulus onset for each trial, and spectral decomposition was performed across the 0.5-100 Hz frequency range, excluding 58–61 Hz to suppress residual line noise. The multitaper method provides robust spectral estimates by averaging across multiple orthogonal tapers.

For analysis, epochs were extracted from-0.3 to 0.7 seconds relative to each pinprick stimulus onset. Time-frequency representations (TFRs) of the oscillatory power (1 to 45 Hz) were computed using real and imaginary signal components of the decomposition and transformed into decibel scale^3, 43^. Baseline correction was performed by subtracting the average pre-stimulus power (-0.3 to 0 s) for each trial per frequency. Corrected dB power responses were then averaged across trials to yield subject-and stimulus condition-specific TFRs for each of the four regions of interest (bilateral mOFC and dlPFC). Each resulting TFR was stored as a time (T) × frequency (F) matrix, downsampled to approximately 37 ms resolution (T = 27, F = 45).

### Statistical Modeling for cLBP Cohort

We modeled baseline-corrected TFRs in the 0 to 0.7 second post-stimulus window as 2D functional outcomes (time × frequency) using a Bayesian mixed-effects framework^44, 45^. For each ROI, we jointly modeled stimulus-evoked responses across four conditions (Back 32 mN, Back 256 mN, Hand 32 mN, Hand 256 mN). The model included fixed effects for group (Widespread, Localized, No Pain), age, sex, and PROMIS pain intensity, and random intercepts for subjects, as well as subject-by-condition random effects, to account for within-subject residual correlation. To represent 2D TFR responses, we used a bivariate cubic B-spline basis (5 × 10 dimensions for time and frequency). Fixed effects were projected onto a rank-1 representation using an automatic rank selection procedure. Covariate coefficients were given zero-mean normal priors with hierarchical shrinkage priors on their variances. The fixed effects’ basis was modeled as an outer product of unit-norm vectors with uniform priors on the unit sphere.

Random effect coefficients received independent zero-mean normal priors with Inverse-Gamma hyperpriors on variances. Measurement error variances had Jeffreys’ priors. We used 1500 burn-in and 500 post-burn-in MCMC iterations. All EEG preprocessing and modeling steps were conducted using MNE-Python, R (including rhdf5 and ggplot2), and custom analysis pipelines developed for this project.

### Transfer Learning for CP Cohort in Predicting Treatment Response

The limited sample size of the CP cohort (n = 18) constrained our ability to directly infer reliable signatures of widespread hyperalgesia within the CP group alone. To address this, we first modeled TFR responses in the cLBP cohort (n = 67) to identify frontal cortical signatures (mOFC and dlPFC) associated with widespread hyperalgesia. This multi-subject TFR analysis in the cLBP cohort informed the development of an EEG-based biomarker of widespread hyperalgesia, which was then applied to the CP cohort to assess its generalizability and predictive value. Specifically, we projected each CP participant’s TFR responses onto the cLBP-derived oscillatory signatures and examined whether these projections distinguished pain phenotypes (widespread, segmental, or no hyperalgesia) and predicted treatment (ERP) response in CP. This cross-condition application in developing predictive models represents a transfer learning framework^46, 47^, leveraging statistical features learned in one population (cLBP) to refine pain phenotyping in another (CP) and use these features to help develop clinical prediction models in CP.

Group differences in biosignature projection scores among CP participants were evaluated using one-way analysis of variance (ANOVA), followed by Tukey’s honest significant difference (HSD) tests for post-hoc comparisons. To predict 3-month ERP response in CP participants, we fit a linear regression model using the cLBP-derived mOFC biosignature, the P-QST-derived hyperalgesia phenotype (widespread vs. segmental/no hyperalgesia) and their interaction. Model performance was compared to two alternative models based solely on P-QST phenotypes (the widespread hyperalgesia model and the three-group phenotype model), with respect to the (adjusted) R² and leave-one-out cross-validation (LOOCV) area under the receiver operating characteristic curve (AUC).

## Results

### A subset of patients in the cLBP and CP cohorts report widespread hyperalgesia

In the cLBP cohort, we recorded stimulus-evoked EEG with mechanical pinprick stimuli of 32 mN and 256 mN to the low back (diseased site) and hand (disease free site); pain-free controls underwent the same protocol. **Table 1** reports baseline participant information for the cLBP cohort and pain-free controls (n=67), including age, sex, and average pain intensity in the prior week, stratified by identified pain phenotype: no pain (control), localized pain, and widespread hyperalgesia (see **Methods**)^38^. The participants who experienced increased pain at an anatomically distant, healthy site (i.e., Widespread Hyperalgesia) comprised about 29% of all the cLBP participants, compatible with previous literature on the incidence of widespread hyperalgesia in chronic pain conditions^1, 3, 4, 48^.

**Table 1:**
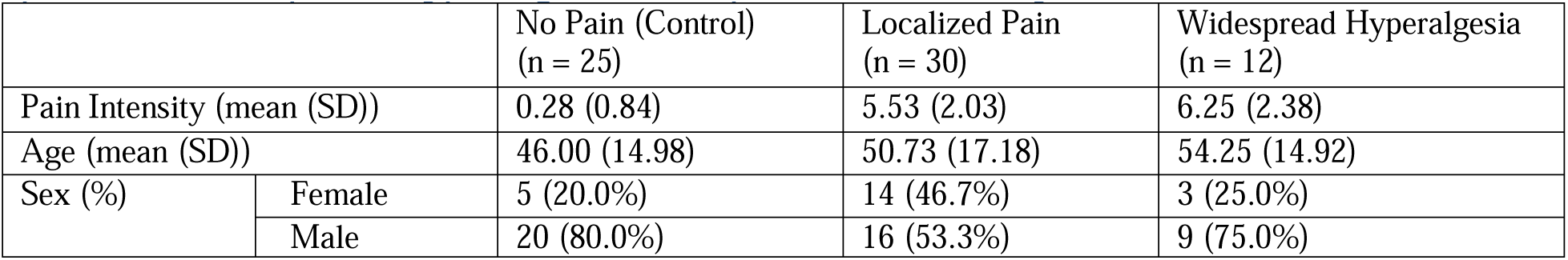
Chronic low back pain participants show different pain phenotypes (localized pain and widespread hyperalgesia) compared with healthy controls.

|  |  | No Pain (Control)<br>(n = 25) | Localized Pain<br>(n = 30) | Widespread Hyperalgesia<br>(n = 12) |
| --- | --- | --- | --- | --- |
| Pain Intensity (mean (SD)) |  | 0.28 (0.84) | 5.53 (2.03) | 6.25 (2.38) |
| Age (mean (SD)) |  | 46.00 (14.98) | 50.73 (17.18) | 54.25 (14.92) |
| Sex (%) | Female | 5 (20.0%) | 14 (46.7%) | 3 (25.0%) |
|  | Male | 20 (80.0%) | 16 (53.3%) | 9 (75.0%) |

**Table 2** reports baseline participant information for the CP cohort (n=18), including age, sex and pain intensity (BPI pain severity subscale, averaged across the four items: Pain Worst, Pain Average, Pain Least, Pain Now), stratified by pain phenotype derived from a standardized pancreatic-specific QST (P-QST) protocol (see **Methods**)^8, 37^. Twelve out of 18 patients were identified as having hyperalgesia, including 8 with widespread hyperalgesia and 4 with localized (i.e., segmental) hyperalgesia.

**Table 2:** Chronic pancreatitis patients show different pain phenotypes.

|  |  | No Hyperalgesia<br>(n = 6) | Segmental Hyperalgesia<br>(n = 4) | Widespread<br>Hyperalgesia (n = 8) |
| --- | --- | --- | --- | --- |
| BPI baseline (mean (SD)) |  | 4.29 (2.58) | 3.38 (2.26) | 3.84 (1.41) |
| Age (mean (SD)) |  | 61.83 (17.37) | 53.25 (7.37) | 52.50 (17.55) |
| Sex (%) | Female | 0 (0.0) | 2 (50.0) | 4 (50.0) |
|  | Male | 6 (100.0) | 2 (50.0) | 4 (50.0) |

### Low-frequency oscillatory changes in the mOFC are associated with widespread hyperalgesia in cLBP participants

We focused our inquiry on the mOFC and dlPFC regions given their roles in chronic pain^6, 10, 14^. After source localization, time-and frequency-resolved oscillatory activity in the mOFC and dlPFC was extracted and analyzed (**Supplementary Figure S1**). To probe nociceptive processing, we applied two different, weighted stimuli (32 mN, non-noxious, and 256 mN, noxious) to back (the site of disease in cLBP participants) and hand (disease-free site). **Supplementary Figure S2** shows time-frequency representations (TFRs) for each pain phenotype group, averaged across participants for the left and right mOFC.

We modeled how pain phenotype groups (widespread hyperalgesia, localized pain, no pain) influenced EEG time-frequency responses to noxious stimulation in the mOFC and dlPFC, adjusting for age, sex, and pain intensity (**Figure 2**). In the left mOFC (**Figure 2**, first row), the estimated 2D basis for the evoked oscillatory activity in the time-frequency domain (first row, left panel) exhibited a characteristic pattern approximately in the 0.4-0.7 second post-stimulus window, with relatively elevated power in the delta/theta (1-7 Hz) range and suppressed power in the alpha (8-12 Hz) range. In this basis, participants with widespread hyperalgesia showed significantly greater stimulus-evoked oscillatory activity than those with localized pain under high-intensity stimulation at the affected site (Back 256 mN), with an estimated group difference of 9.09 (95% CI: [5.61, 12.69]) (**Figure 2**, top row, right panel) (this coefficient, when multiplied by the basis in the left panel, represents the group difference in the time-frequency domain in oscillatory power, expressed in decibels). Similar data-driven time-frequency basis patterns were observed in the right mOFC (**Figure 2**, second row, left panel). However, no statistically significant phenotype group differences were observed in both the left and right dlPFC (**Figure 2**, third and fourth rows).

**Figure 2.**
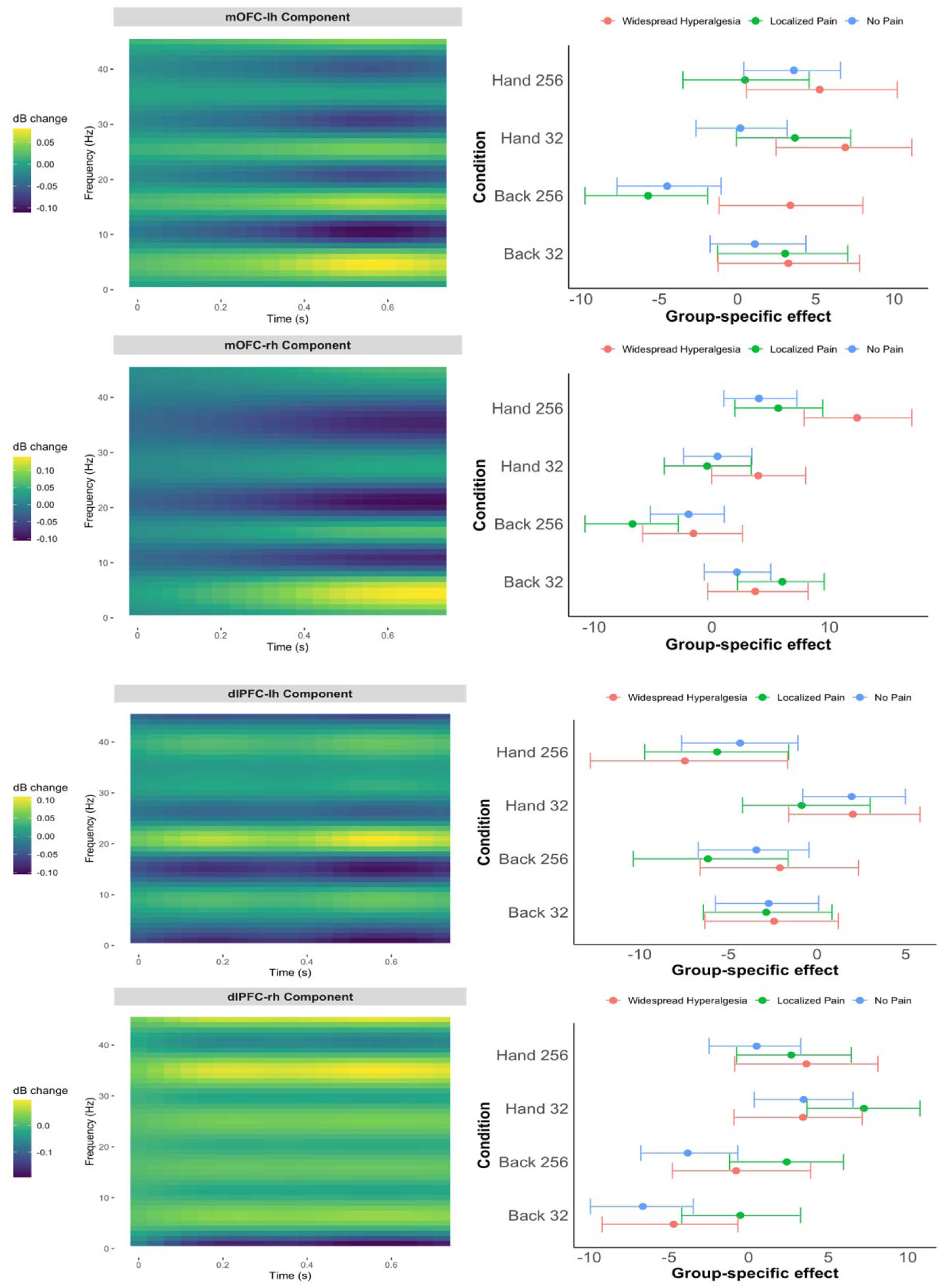
Pain phenotype effects on time-frequency responses in the mOFC (top two rows) and dlPFC (bottom two rows) based on cLBP data (n=67). Left panels display the data-driven time-frequency basis for the power; right panels show posterior summaries of group-by-condition effects projected onto that basis (means and 90% credible intervals), adjusted for pain intensity, age, and sex. Results are shown by stimulus condition (Back 32 mN, Back 256 mN, Hand 32 mN, Hand 256 mN) and laterality (top: left hemisphere (lh); bottom: right hemisphere (rh)).

### Development of an EEG biosignature for widespread hyperalgesia

The observed differences in low-frequency (delta, theta and alpha) oscillatory activity of the mOFC between participants with widespread versus localized pain suggest a candidate biosignature for widespread hyperalgesia. We defined an EEG-based biosignature of widespread hyperalgesia (versus localized pain) as a projection score that quantifies the expression of this characteristic mOFC oscillatory pattern. Specifically, the biosignature was constructed as a weighted sum of the oscillatory power in the left and right mOFC during the 0.4–0.7 second post-stimulus window, under high-intensity stimulation at both affected and unaffected sites and low intensity stimulation at unaffected site. The power in the 1–7 Hz (delta/theta) range was assigned a weight of +1, and the power in the 8–12 Hz (alpha) range was assigned a weight of-1, such that higher scores reflect stronger expression of the widespread hyperalgesia signature—characterized by increased delta/theta and suppressed alpha activity. This time-frequency projection pattern is illustrated in **Figure 3**.

**Figure 3.**
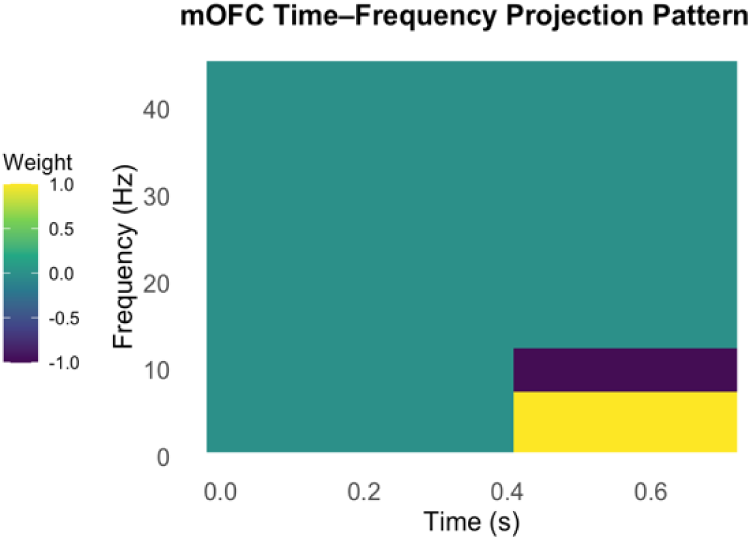
Time-frequency projection pattern for the mOFC used in biosignature. The pattern highlights increased delta/theta (1–7 Hz) (weighted by +1) and decreased alpha (8–12 Hz) (weighted by - 1) oscillatory power during the 0.4–0.7 second post-stimulus window. This weighting scheme was applied to each participant’s mOFC time-frequency response (left and right hemispheres, averaged) to compute a scalar biosignature score reflecting expression of the widespread hyperalgesia profile.

### Testing the biosignature for widespread hyperalgesia in a new chronic pain (CP) cohort

Analogous to the cLBP cohort, in the CP cohort, we applied two levels of mechanical stimulation (32 mN, non-noxious; 512 mN, noxious) to both the upper abdomen (disease-affected site in CP participants) and the forearm (disease-free site). **Supplementary Figure S3** presents group-averaged time-frequency EEG responses for the CP cohort, grouped by pain phenotype (Widespread, Segmental, No Hyperalgesia), where the widespread hyperalgesia group exhibited greater delta/theta (1-7 Hz) power under high-intensity stimulation at the affected site (Abdomen 512 mN) compared to the segmental hyperalgesia group. In contrast, the segmental hyperalgesia group showed relatively reduced delta/theta activity at the unaffected site (Forearm 512 mN) compared to the widespread hyperalgesia group.

To assess the generalizability of mOFC oscillatory features as a marker of widespread hyperalgesia, we applied the cLBP-derived EEG biosignature to participants with CP. **Figure 4** (top row) displays the mean (± standard error of the mean; SEM) projection scores onto the EEG biosignature for the CP cohort (n = 18), grouped by pain phenotype. ANOVA indicated statistically significant group differences (left: *p* = 0.037; right: *p* = 0.036) in the CP cohort, indicating that projection of the CP participants’ EEG responses onto this cLBP-derived biosignature differentiated the three CP pain phenotypes, with comparable discriminative capacity for both the left and right mOFC scores (**Figure 4**, left and right panels, respectively). Tukey’s HSD post-hoc comparisons indicated higher projection scores in the widespread hyperalgesia group compared to the segmental hyperalgesia group (left: *p* = 0.048; right: *p* = 0.036).

**Figure 4.**
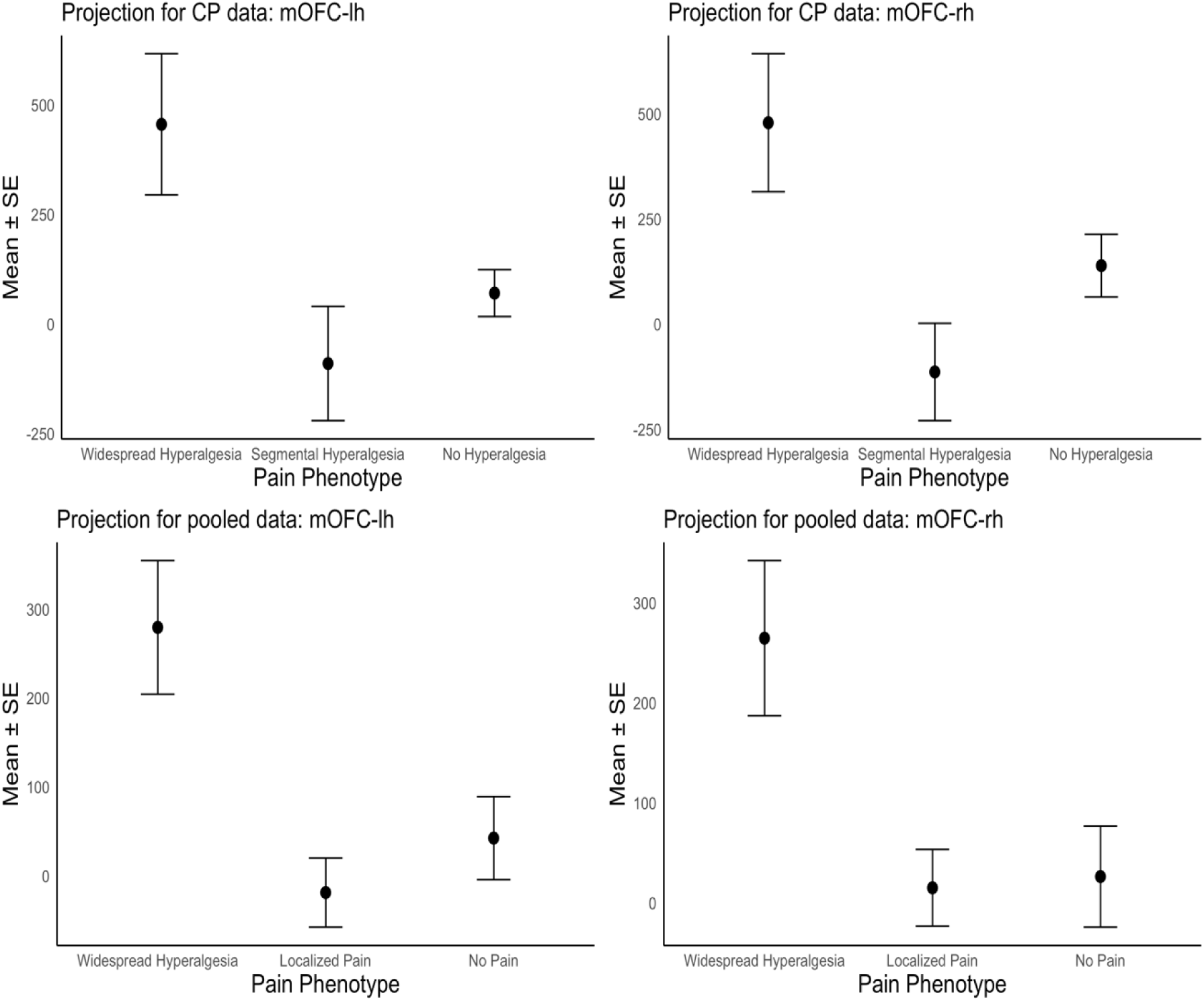
**Top panels**: **Projection scores onto the mOFC biosignature in the CP sample (n=18), by 3 pain phenotype groups**, for the left and right mOFC (left and right panels), respectively. Group means ± standard error of the mean (SEM) are shown for CP participants, grouped by pain phenotype (Widespread, Segmental, No Hyperalgesia). The mOFC projection scores successfully differentiated pain phenotypes, with consistent patterns observed across the both hemispheres. **Bottom panels**: **Projection scores onto the mOFC biosignature based on the pooled (cLBP + CP) sample (n=85), by 3 pain phenotype groups**, for the left and right mOFC (left and right panels), respectively. Group means ± SEM for participants in the pooled sample, grouped by pain phenotype (Widespread Hyperalgesia, Localized Pain, No Pain). The mOFC projection scores successfully distinguished pain phenotypes, with consistent patterns observed across the both hemispheres.

These findings support the applicability of the cLBP-derived biosignature in distinguishing widespread hyperalgesia in CP, suggesting that low-frequency oscillatory changes in the mOFC may reflect a shared neurophysiological mechanism.

To further assess the robustness of the mOFC signature of widespread hyperalgesia, we pooled data from both the cLBP (n=67) and CP (n=18) cohorts (total n = 85) and repeated the analyses conducted above. For this pooled analysis, we harmonized the two cohorts using widespread hyperalgesia phenotype as a common marker. The “Localized Pain” group refers to participants with either localized hypersensitivity (i.e., “Segmental Hyperalgesia” in CP) or no measurable hyperalgesia (i.e., “No Hyperalgesia” in CP and “Localized Pain” in cLBP). Consistent with findings from the CP cohort alone, projection of participants’ responses onto the mOFC biosignature distinguished the three pain phenotype groups, with similar discriminative capacity for the left and right mOFC scores (**Figure 4**, bottom panels). In the pooled dataset, one-way ANOVA confirmed significant group differences (left: p < 0.001; right: p = 0.003; **Figure 4**, bottom panels), and Tukey’s HSD post-hoc comparisons indicated significantly higher projection scores for the Widespread Hyperalgesia group compared to the Localized Pain group (left: p < 0.001; right: p = 0.004).

Further, we repeated the analysis presented in **Figure 2**, using the pooled data (n = 85), adjusting for pain intensity, age, sex, and cohort. As in **Figure 2**, **Figure 5** displays the data-driven time-frequency basis (left panels) alongside posterior summaries of group-by-condition effects on the evoked oscillatory power projected onto that basis (right panels). Consistent with the cLBP-only analysis, the pooled data revealed a prominent oscillatory pattern in the mOFC, characterized by elevated delta/theta (1–7 Hz) and reduced alpha (8–12 Hz) activity approximately in the 0.4–0.7 second post-stimulus window (**Figure 5**, top left panels), that differentiated widespread hyperalgesia from localized pain (**Figure 5**, top right panels). These findings reinforce the specificity of the mOFC oscillatory signature for widespread hyperalgesia and support its transdiagnostic relevance.

**Figure 5.**
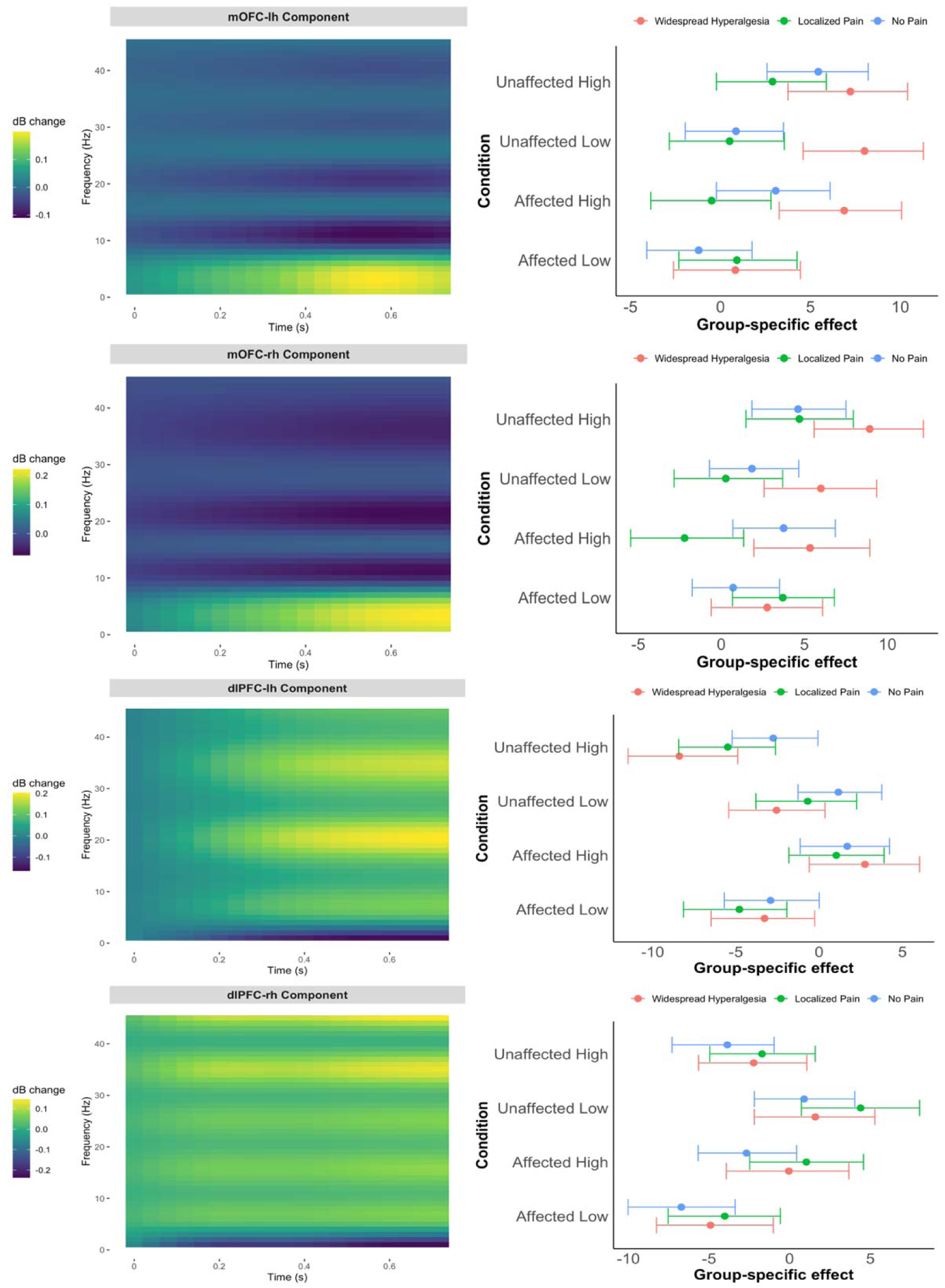
Pain phenotype effects on time-frequency responses in the mOFC and dlPFC based on pooled data (cLBP and CP; n=85). Left panels display the data-driven time-frequency basis; right panels show posterior summaries of group-by-condition effects projected onto that basis (means and 90% credible intervals), adjusted for pain intensity, age, sex and cohort. Results are shown by stimulus condition (Affected/Unaffected site and High/Low intensity) and laterality (top: left hemisphere; bottom: right hemisphere). In the pooled analysis, Localized Pain refers to regional pain with either localized hypersensitivity or no measurable hyperalgesia.

### Prediction of treatment response in CP

A subset of participants in the CP cohort underwent ERP for the treatment of pancreatic ductal obstruction (*n* = 11). ERP is known as a treatment that specifically targets the peripheral mechanisms of pain, but it is thought to have limited efficacy for widespread hyperalgesia that may have central mechanisms, as has been shown for peripherally targeted interventions in other pain conditions associated with widespread hyperalgesia^12, 49^. We assessed pain at 3-months after a technically successful ERP treatment. Among participants without widespread hyperalgesia (n=6), 3 were classified as treatment responders (characterized as ≥30% reduction in BPI pain severity at 3-months relative to baseline BPI) (**Figure 6**, left panel). In contrast, no responders were observed among those with widespread hyperalgesia (n=5).

**Figure 6.**
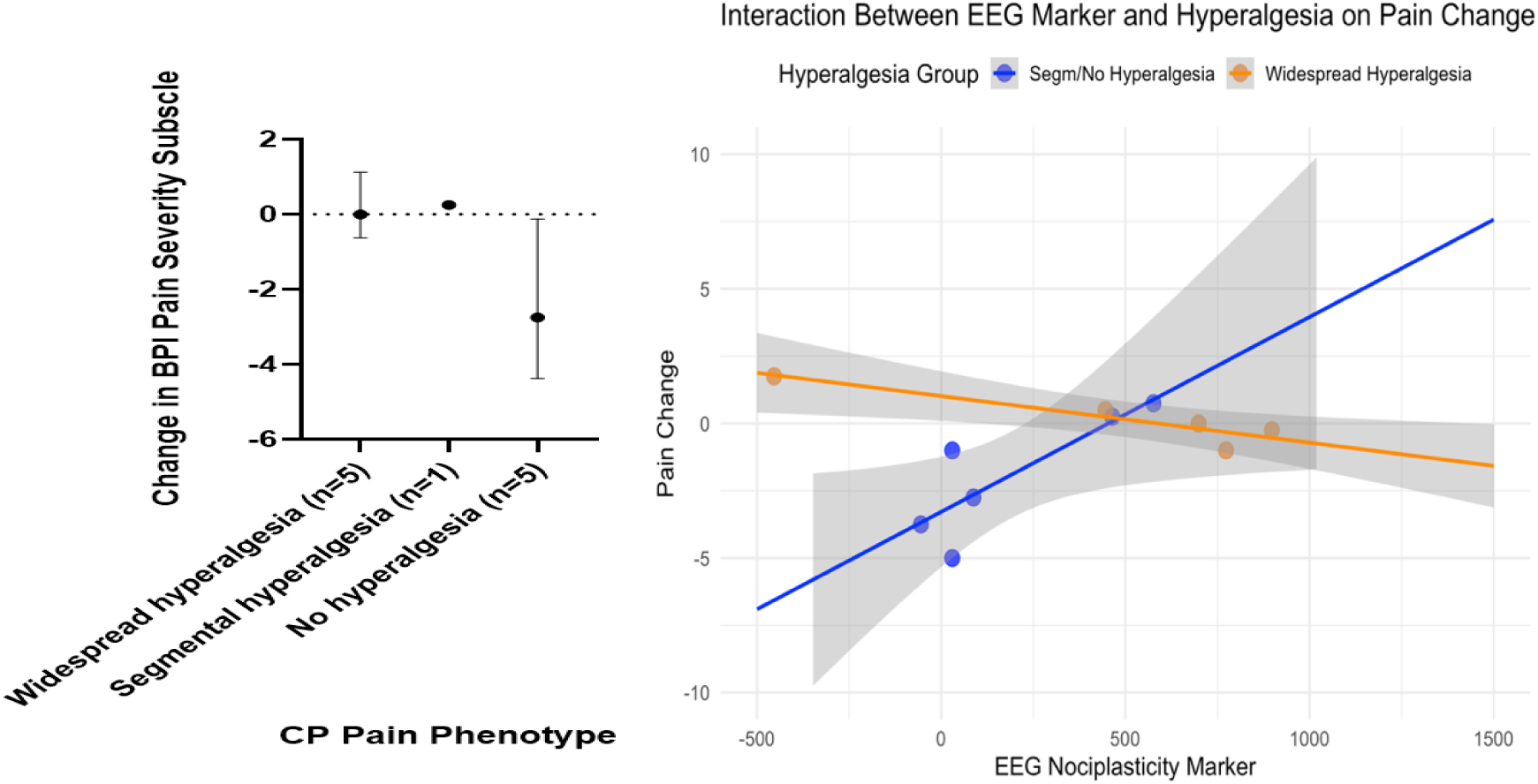
**Left panel**: **Change in Brief Pain Inventory (BPI) pain severity subscale at 3 months after a technically successful ERP, stratified by pain phenotypes**. Negative values indicate improvement in pain. Values are median and interquartile range (IQR). Overall, participants with widespread or segmental hyperalgesia showed minimal or no responses while participants with no hyperalgesia exhibited better responses, albeit with greater variability.

### Right panel: Post-treatment pain change

(negative values indicate pain improvement) vs. baseline EEG biomarker scores. Linear regression lines are shown for participants with and without widespread hyperalgesia. The interaction model (EEG × widespread hyperalgesia) that incorporated the EEG marker outperformed models based on the widespread hyperalgesia indicator alone or the 3-level pain phenotypes ( = 71% vs. 21% and 27%) and achieved LOOCV AUC = 88% (vs. 63% and 71%).

We next fit a linear regression model using the mOFC-based EEG biosignature, P-QST-derived hyperalgesia phenotype (widespread hyperalgesia vs. segmental or no hyperalgesia), and their interaction to predict 3-month treatment response in CP participants (**Figure 6**, right panel). We compared this multimodal model based on both the EEG biosignature and P-QST-derived phenotype, against two other models for predicting treatment response in CP that are based solely on P-QST results. Our model that incorporated the EEG biosignature (R^2^ = 80%; = 71%) substantially outperform these two behavior-based models in terms of the (adjusted) R^2^: one using only P-QST-based binary widespread hyperalgesia phenotype (R^2^ = 29%; = 21%), and another using P-QST-based three-level phenotype (widespread, segmental, and no hyperalgesia) categories (R^2^ = 42%; R^2^*_adjusted_*= 27%). Among participants without widespread hyperalgesia (i.e., the Segmental/No hyperalgesia group in **Figure 6**, right panel), lower EEG biosignature scores were correlated with improvement in pain, suggesting that combining the mOFC-based biomarker with standard P-QST results can refine phenotyping and produce better prediction for the success of peripherally driven therapies.

Leave-one-out cross-validation (LOOCV) further supported the added predictive value of the EEG biosignature. The interaction model that incorporated the EEG biosignature achieved a LOOCV

AUC exceeding 85% (AUC = 88%; accuracy = 82%; sensitivity = 67%; specificity=88%), outperforming the widespread hyperalgesia-only model (LOOCV AUC = 63%; accuracy = 64%; sensitivity = 67%; specificity=63%) and the 3-level categorical phenotype model (LOOCV AUC = 71%; accuracy = 70%; sensitivity = 67%; specificity=71%).

Together, these results indicate that a shared oscillatory brain signature of widespread hyperalgesia may exist across chronic pain conditions (cLBP and CP), and that these cortical nociceptive responses can be leveraged as a biosignature for risk stratification and prediction of treatment response.

## Discussion

In this study, we found that cLBP participants with widespread hyperalgesia, compared to localized or no pain, exhibited a characteristic pattern of increased delta/theta coupled with decreased alpha oscillatory power in the mOFC during noxious pinprick stimulation. Based on these findings, we derived a biosignature of widespread hyperalgesia and found that this cLBP-derived marker can be effectively applied to CP participants, a distinct chronic patient population, for pain phenotype differentiation.

Notably, CP participants with widespread hyperalgesia, compared to segmental or no hyperalgesia, showed an elevated expression of the mOFC delta–theta oscillatory marker with alpha suppression. Despite differences in peripheral pathology between cLBP and CP, our results suggest a potential common central mechanism for widespread hyperalgesia.

### Role for mOFC in widespread hyperalgesia

The involvement of mOFC in widespread hyperalgesia is compatible with its role in affective and cognitive pain modulation. This region is known to receive signals from and in turn project to multiple cortical areas. The oscillatory changes observed in mOFC may thus reflect reduced cortical inhibition (increased oscillatory power in slow-wave bands), consistent with prior literature^26, 27, 33^. For example, increased theta activity has been reported in participants with various chronic pain conditions, both in resting state recordings as well as evoked EEG potentials^28, 50–55^. In our cLBP cohort, painful stimulation at the site of injury (lower back) evoked significantly greater theta-band responses, with a relative alpha suppression, in participants with widespread hyperalgesia compared to localized pain or pain-free controls. Compatible with our findings here, a recent study in chronic pain participants with chronic intracranial recordings showed that changes in oscillatory activities in the OFC can function as a biomarker for chronic pain^14^.

Other studies have shown that mOFC has prominent roles in reward processing as well as placebo analgesia^56, 57^.

Placing our specific findings on nociceptive processing in widespread hyperalgesia within the current literature, a model emerges in which the mOFC acts as a critical hub in central regulation of pain, by integrating cognitive-affective information with sensory-nociceptive signals. As a result, when the activity of mOFC is dysregulated, there is decreased top-down regulation, resulting in widespread hyperalgesia and pain amplification.

### Mechanism-based biosignature for widespread hyperalgesia

Methodologically, we leveraged projection-based biomarker construction to develop an EEG index of widespread hyperalgesia. To assess generalizability, we projected CP participants’ EEG responses onto the cLBP-derived oscillatory signature. This cross-condition application suggests the promise of EEG-derived markers in identifying central pain amplification. Rather than relying on a single frequency band, our biosignature incorporates the multi-band oscillatory pattern (delta/theta increase with alpha decrease in mOFC) as a composite feature. We achieved this by identifying the mOFC time-frequency representation (TFR) that best differentiated the widespread hyperalgesia subgroup and then projecting individual EEG data onto this signature to yield a scalar index for each participant, that can be compared across individuals.

In both the cLBP and CP cohorts, biomarker values showed a consistent difference between widespread hyperalgesia vs. segmental hyperalgesia or localized pain. Our biosignature, defined as increased delta/theta and decreased alpha power in the mOFC during the 0.4-0.7 second post-stimulus window, appears to reflect the extent of central affective-cognitive engagement in pain processing. Participants with widespread hyperalgesia showed higher biosignature values, consistent with greater centralized sensitization and diminished top-down inhibitory control. In contrast, participants with localized hyperalgesia exhibited lower biosignature values, suggesting a pain phenotype with less mOFC-mediated central component, with pain dominated by peripheral mechanisms. These findings support the utility of mOFC-based oscillatory activities as a marker that represents the spectrum of widespread hyperalgesia.

### Predictive value for an EEG biosignature in chronic pain syndromes

Our EEG biosignature differentiated pain phenotypes. More importantly, it added predictive value for 3-month treatment outcomes in CP participants, outperforming P-QST-based signatures of widespread hyperalgesia. Higher scores of our biosignature were associated with poor treatment response among participants without hyperalgesia, suggesting its potential for early identification of at-risk individuals and personalization of therapy.

Both cLBP and CP are clinically heterogeneous. In some patients, pain is dominantly driven by ongoing peripheral nociceptive, inflammatory or neuropathic processes, while others develop a disproportionate or widespread pain sensitivity that may have a strong component of central amplification or nociplastic pain. Differentiation between peripherally vs. centrally-driven pain is clinically important, as it can inform treatment selections. Participants who have pronounced central amplification may not respond to therapies targeting peripheral processes. Identifying individuals with CP associated with widespread hyperalgesia will have important impact on their treatment as these individuals would likely not benefit from pancreatic targeted interventions such as ERP or surgery. Prior studies have explored the use of QST to identify participants more likely to benefit from peripheral interventions like ERP, such as those without central sensitization^58, 59^. Our results here indicate that addition of EEG biosignature to P-QST can enhance the predictive accuracy by further explaining the heterogeneous responses to ERP.

### A transfer learning approach to study mechanisms of chronic pain

While this study focused on cLBP and CP, the concept of a shared nociplastic EEG signature may extend to other chronic pain conditions involving central sensitization (e.g., osteoarthritis). Our success to transfer oscillatory signatures that we learned from cLBP to CP suggests potential shared neurophysiological mechanisms of widespread hyperalgesia and underscores the potential use of EEG-based biomarkers for cross-condition phenotyping. This transfer learning approach offers a data-efficient strategy to accelerate biomarker development in underpowered clinical populations.

### Limitations and future directions

Key limitations include the small sample size in the CP cohort (n = 18) that precluded reliable estimation of oscillatory effects of widespread hyperalgesia from the CP cohort alone, and the lack of an independent validation dataset for predictive models. Thus, the EEG biosignature derived here should be prospectively tested as a stratification tool at baseline to track clinical outcomes longitudinally in a larger cohort to further validate its predictive accuracy. Longitudinal studies could also assess how the EEG signature changes with effective treatment and whether it can serve as an objective treatment-induced neuroplasticity measure. Furthermore, our analyses focused on local oscillatory power changes in specific brain regions and did not examine functional connectivity. Incorporating EEG connectivity or graph theoretical metrics in future work could enrich our understanding of how widespread hyperalgesia emerges from distributed brain circuit dysfunction and could yield multi-feature biomarkers that further improve classification accuracy. Another important direction is to integrate EEG with structural and functional neuroimaging to build a more comprehensive biomarker. Structural MRI could be used to see if there are anatomical correlates of the EEG signature (e.g., cortical thinning in those with high mOFC theta power)^60^. Combining EEG and fMRI simultaneously can also leverage the strengths of each modality to more precisely characterize the nociplastic signature^61^.

## Summary

We identified an mOFC-based EEG signature of widespread hyperalgesia that may serve as a mechanistic marker of pain amplification across two different chronic pain syndromes. Our results suggest potential shared mechanisms among widespread hyperalgesia and the possibility of mechanistically driven biomarkers that can predict treatment success to advance precision pain medicine.

## Data Availability

All code and processed data used in this study are available at https://github.com/syhyunpark/mOFC-EEG-biosignature-hyperalgesia. The repository includes scripts and data necessary to reproduce all analyses, figures, and tables presented in this preprint. Data from the CP cohort will be deposited in the NIMH Data Archive as part of our funded study.

## Author Contributions

Conceptualization: HGP, ZSC, TAG, JW, LVD; Methodology: HGP, XJ, TAG, JW, LVD; Investigation: MMR, RRW, TAG, JW, LVD; Visualization: HGP, XJ, MMR, GK, LVD; Supervision: HGP, TAG, JW, LVD; Writing—original draft: HGP, JW, LVD; Writing—review & editing: All

## Supporting Information for

**Figure S1.**
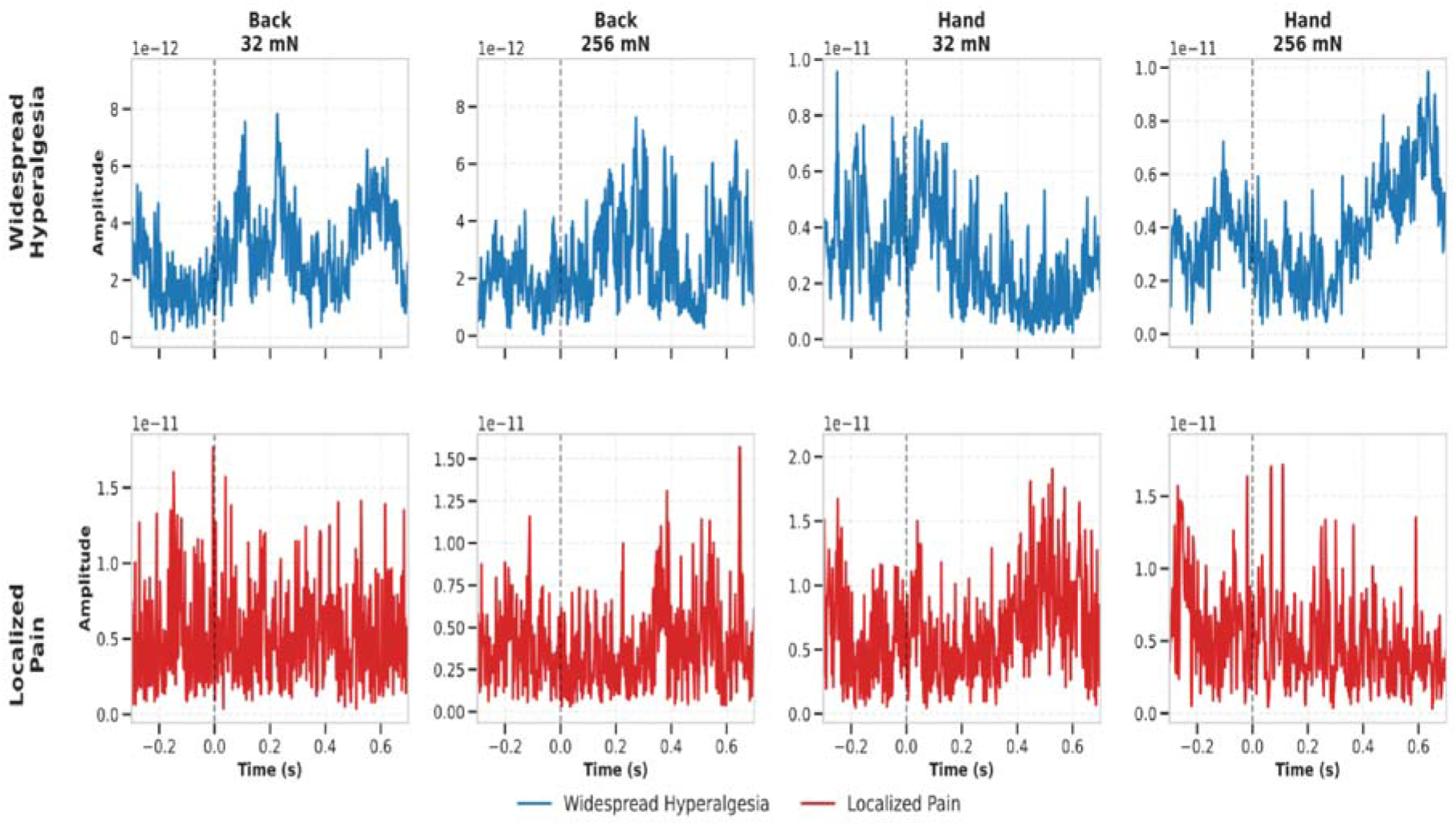
**Source-localized EEG traces from the left mOFC in representative cLBP participants. Top row**: Source-localized EEG traces from the mOFC (left hemisphere) for a representative participant with widespread hyperalgesia, shown across four stimulus conditions (Back 32 mN, Back 256 mN, Hand 32 mN, Hand 256 mN). **Bottom row**: Traces from a representative participant with localized pain, under the same four stimulus conditions. Epochs were extracted from-0.3 to 0.7 seconds relative to each pinprick stimulus onset. Time-frequency representations (TFRs) of power (1 to 45 Hz) were computed using real and imaginary signal components of the decomposition and transformed into decibel scale. Baseline correction was performed by subtracting the average pre-stimulus power (-0.3 to 0 s) for each trial per frequency. Corrected dB power responses were then averaged across trials per condition to yield subject-and stimulus condition-specific TFRs for each of the four regions of interest (bilateral mOFC and dlPFC).

**Figure S2.**
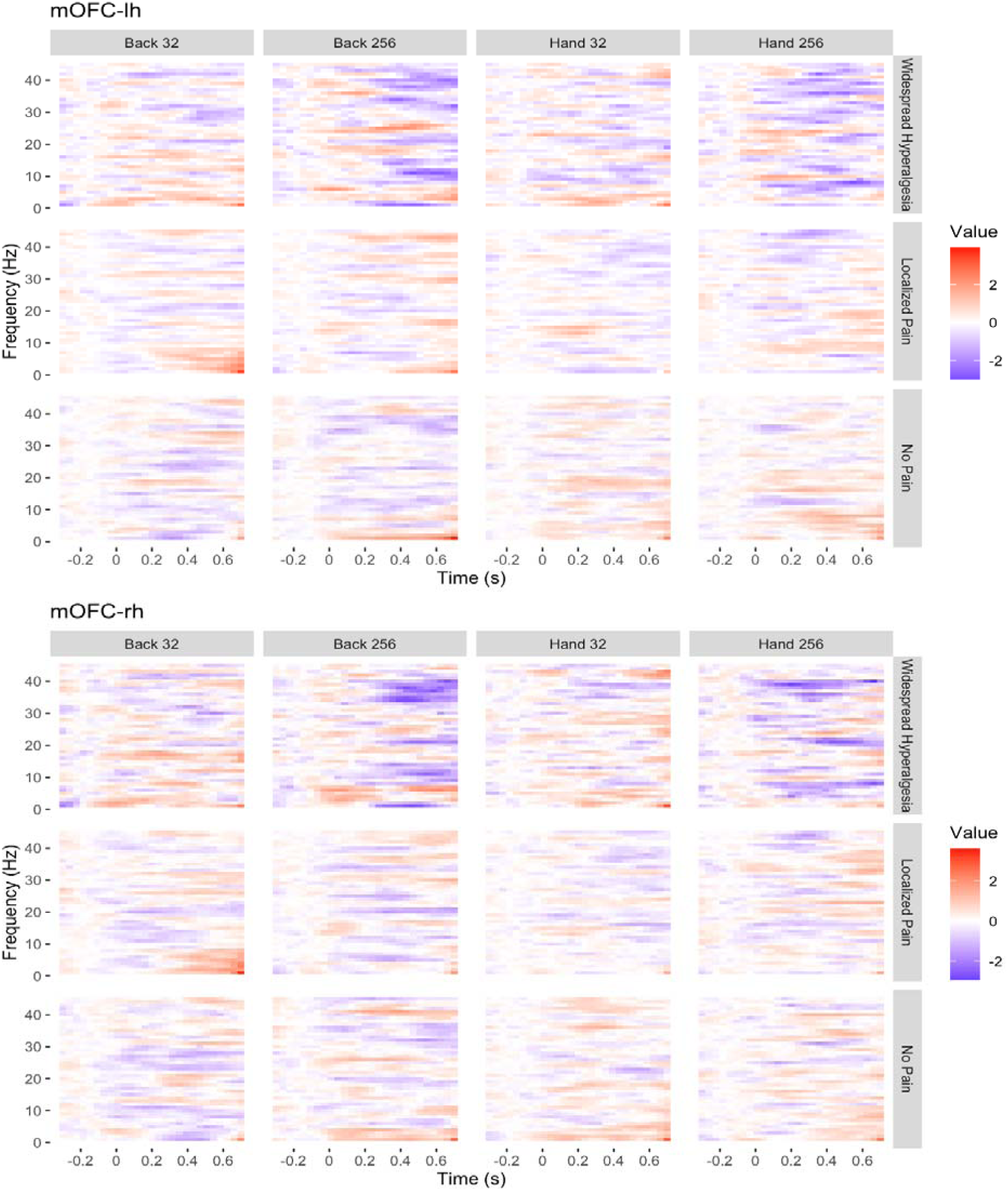
**Time-frequency EEG responses in the mOFC by stimulus condition and pain phenotype in the chronic low back pain (cLBP) cohort (n = 67)**. Group-and stimulus-specific averaged time-frequency representations (TFRs) for the cLBP cohort, averaged across participants within each pain phenotype group (Widespread Hyperalgesia, Localized Pain, No Pain). Top panels show responses from the left mOFC; bottom panels from the right mOFC. Each panel displays baseline-corrected decibel power from-0.3 to 0.7 seconds post-stimulus across four conditions: Back 32 mN, Back 512 mN, Hand 32 mN, and Hand 512 mN.

**Figure S3.**
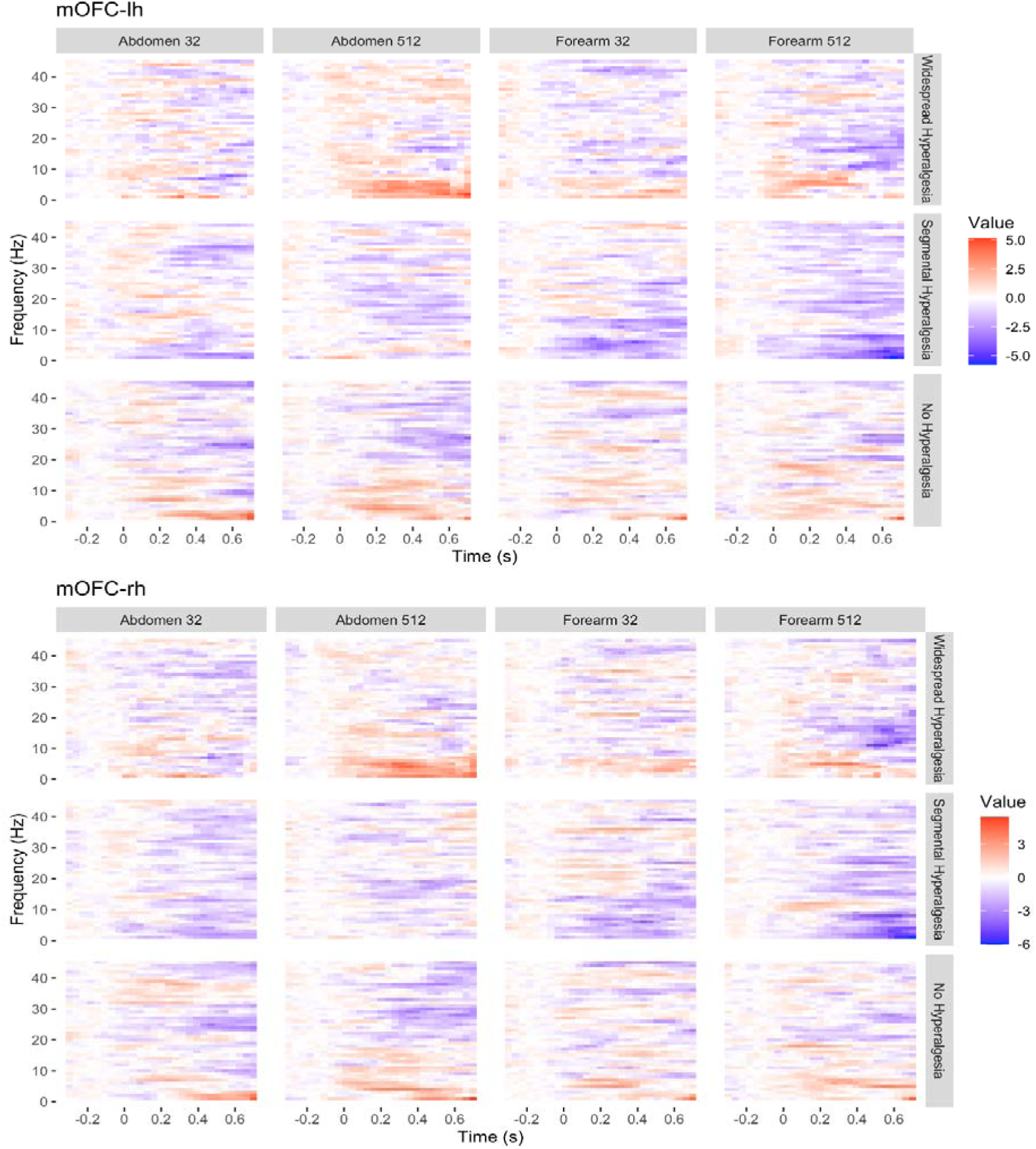
Time-frequency EEG oscillatory responses in the mOFC by stimulus condition and pain phenotype in the chronic pancreatitis (CP) cohort (n = 18). Group-averaged time-frequency representations (TFRs) are shown for each pain phenotype group (Widespread, Segmental, No Hyperalgesia), stratified by stimulation site and intensity: Abdomen 32 mN, Abdomen 512 mN, Forearm 32 mN, and Forearm 512 mN. Each panel displays baseline-corrected power (in decibels) from –0.3 to 0.7 seconds post-stimulus. Top panels reflect responses from the left mOFC; bottom panels from the right mOFC. The widespread hyperalgesia group (top row) exhibited elevated delta/theta (1–7 Hz) power under high-intensity stimulation at the affected site (Abdomen 512 mN), compared to the segmental hyperalgesia group (middle row). The segmental hyperalgesia group showed relatively reduced delta/theta activity at the unaffected site (Forearm 512 mN), suggesting differential neural processing of noxious input across phenotypes.

